# Mechanisms of central brain atrophy in multiple sclerosis

**DOI:** 10.1101/2022.03.28.22273015

**Authors:** Samuel Klistorner, Michael H Barnett, Stuart L Graham, Con Yiannikas, John Parratt, Alexander Klistorner

## Abstract

**Background and objectives:** The measurement of longitudinal change in ventricular volume has been suggested as an accurate and reliable surrogate of central brain atrophy (CBA), potentially applicable to the everyday management of patient with multiple sclerosis (MS). To better understand mechanisms underlying central brain atrophy in RRMS patients we investigated the contribution of inflammatory activity in different lesion compartments to the enlargement of ventricular CSF volume. In addition, we investigated the role of the severity of lesional tissue damage in CBA progression.

**Methods:** Pre- and post-gadolinium 3D-T1, 3D fluid-attenuated inversion recovery (FLAIR) and diffusion tensor images were acquired from 50 patients with relapsing MS. Lesional activity between baseline and 48 months was analysed on FLAIR images using custom-build software, which independently segmented expanding part of the chronic lesions, new confluent lesions and new free-standing lesions. The degree of lesional tissue damage was assessed by change in Mean Diffusivity (MD). Volumetric change of lateral ventricles was used as a measure of central brain atrophy.

**Results:** During follow-up ventricles expanded on average by 12.6+/-13.7%. There was significant increase of total lesion volume, 69.3% of which was due to expansion of chronic lesions and 30.7%-to new (confluent and free-standing) lesional activity. There was high degree of correlation between volume of combined lesional activity and CBA (r^2^=0.67), which became considerably stronger when lesion volume was adjusted by the degree of tissue damage severity (r^2^=0.81). Linear regression analysis explained 90% of CBA variability and revealed that chronic lesion expansion was by far the largest contributor to ventricular enlargement (Standardized Coefficient Beta 0.68 (p<0.001) for expansion of chronic lesions compared to 0.29 (p=<0.001) for confluent lesions and 0.23 (p=0.001) for free-standing new lesions). Age and baseline ventricular volume also provided significant input to the model.

**Discussion:** Our data suggest that central brain atrophy is almost entirely explained by the combination of the volume and severity of lesional tissue activity. Furthermore, the expansion of chronic lesions plays a central role in this process.

## Introduction

Whole-brain atrophy has been extensively used as a measure of neurodegeneration in multiple sclerosis (MS). It predicts worsening of overall disability and cognition when large cohorts of patients are analysed and has been incorporated into newer measures of disease activity such as “NEDA-4” (Alroughani et al., 2016) (Stefano et al., 2014). It also represents an attractive outcome measure for clinical trials of new neuroprotective therapies (Beck & Reich, 2018). There is a growing demand for a personalized approach to MRI measures of MS progression that potentially can be used in clinical practice(Granziera et al., 2021). However, a recent consensus paper on global brain atrophy cautioned against simple “translation of group-based results into actionable, patient-level information”(Sastre-Garriga et al., 2020).

Alternatively, longitudinal measurement of ventricular change has been suggested as an accurate and reliable mean of central brain atrophy (CBA), potentially applicable to everyday patient management(Zivadinov et al., 2016) (M. G. Dwyer et al., 2018) (Mak et al., 2017). Therefore, understanding the biological mechanisms leading to ventricular enlargement is important. While ventricular growth has been related to lesional activity, the contribution of the severity of the lesional damage has not been quantitively examined. Furthermore, better characterization of the relative impact of acute vs chronic (“slow-burning”) inflammation on ventricular expansion in patients with MS is required.

Therefore, to understand mechanisms underlying CBA in relapsing remitting (RRMS) patients we investigated the contribution of both new and ‘slow-burning’ inflammatory activity to enlargement of ventricular CSF volume. In addition, we examined the role of the severity of lesional tissue damage in CBA progression.

## Methods

The study was approved by University of Sydney and Macquarie University Human Research Ethics Committees and followed the tenets of the Declaration of Helsinki. Written informed consent was obtained from all participants.

### Subjects

Fifty consecutive patients with established RRMS, defined according to the revised McDonald 2010 criteria(Polman et al., 2011), were enrolled in a longitudinal study. Patients underwent MRI scans and clinical assessment at 0 months, 12 months and 60 months. The main analysis was performed between 12 months (termed “baseline”) and 60 months (termed “follow-up”), while scans performed at 0 months (termed “pre-study scans”) were used to identify (and exclude) newly developed lesions at the start of the study (i.e. between 0 and 12 months).(S. Klistorner et al., 2021)

### MRI protocol and analysis

MRI was performed using a 3T GE Discovery MR750 scanner (GE Medical Systems, Milwaukee, WI). The following MRI sequences were acquired: Pre- and post-contrast (gadolinium) Sagittal 3D T1, FLAIR CUBE, diffusion weighted MRI. Specific acquisition parameters and MRI image processing are presented in Supplementary material.

JIM 9 software (Xinapse Systems, Essex, UK) was used to segment individual lesions at baseline and follow-up (using co-registered T2 FLAIR images). Lesional activity was analysed using custom-built software that identifies the stable (stationary) component of chronic lesions and additionally segments three separate new lesional compartments: the expanding component of chronic lesions, new confluent lesions and new free-standing lesions, as described previously.(S. A. Klistorner et al., 2020) (Elliott et al., 2019)

The degree of lesional tissue damage was determined by measurement of Mean Diffusivity (MD)(Castriota-scanderbeg et al., 2003) (A. Klistorner et al., 2018). Progressive tissue destruction in each new lesional compartment was calculated independently by measuring the increase of MD between baseline and follow-up timepoints(A. Klistorner et al., 2018) (Wang et al., 2019). Lesion masks for each compartment were adjusted to correct for brain atrophy-related displacement of lesions at follow-up as previously described (S. Klistorner et al., 2021).

To measure the combined effect of brain damage caused by lesional activity of each type, we computed a progressive volume/severity index (PVSI) for each lesional compartment independently by multiplying lesion volume (i.e. the volume of lesion expansion *or* the volume of new confluent lesions *or* the volume of new free-standing lesions) by change of MD in corresponding compartment. The total PVSI was calculated for each patient as the sum of PVSI in each compartment.

Volumetric change of the lateral ventricles was used as a measure of CBA(M. Dwyer et al., 2017) (Kalkers, Vrenken, Uitdehaag, Polman, & Barkhof, 2002). It was calculated by multiplying baseline ventricle volume (as segmented by SIENAX) by the percentage ventricular volume change (as calculated by VIENA, an FSL tool).(Smith et al., 2002) (Vrenken et al., 2014). The volume of whole-brain tissue loss was calculated by multiplying the percentage brain volume change (PBVC) (as determined by SIENA) by baseline brain volume (as segmented by SIENAX).

### Statistics

Statistical analysis was performed using SPSS 22.0 (SPSS, Chicago, IL, USA). Pearson correlation coefficient was used to measure statistical dependence between two numerical variables. For partial correlation, data was adjusted for age, gender, disease duration, baseline lesion volume and ventricular size. P < 0.05 was considered statistically significant. Shapiro–Wilk test was used to test for normal distribution. Comparisons between groups were made using Student *t*-test. Longitudinal changes were assessed using paired two-sample *t*-test. Fisher’s exact test was used for categorical data. Univariate linear regression analysis was performed to estimate the contribution of various new lesional compartments and the severity of tissue damage to brain atrophy.

## Result

Fifty consecutive MS patients who were enrolled in longitudinal study of MS-related axonal loss and who had 5 years follow-up were included in the study. 11 patients were maintained on low-efficacy treatment (injectables, such as interferon and glatiramer acetate, teriflunomide and dimethyl-fumarate)(Samjoo et al., 2021), while 21 patients were receiving high-efficacy drugs (fingolimod, natalizumab, and alemtuzumab)(Samjoo et al., 2021) during the study period. Two patients were treatment-free, while 16 patients changed treatment category between baseline and follow-up visits. Demographic data in presented in Table 1.

**Table 1.**
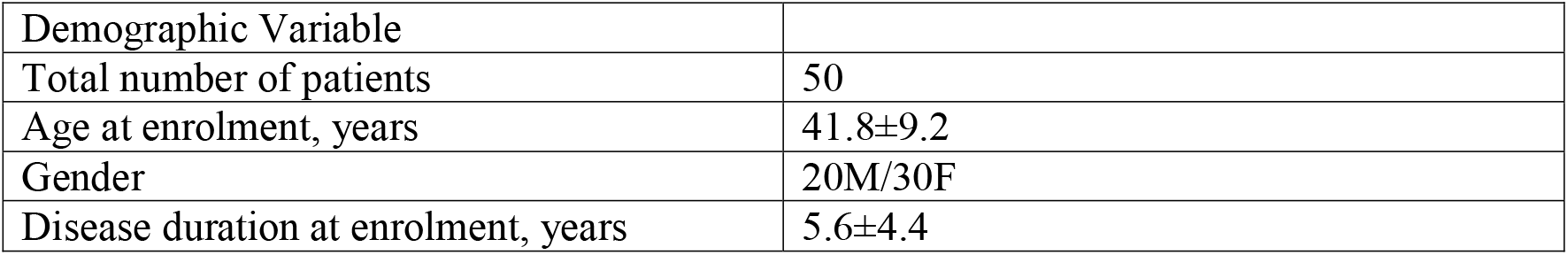
Demographic data.

During follow-up brain volume decreased by 1.55+/-1.2% (as measured by PBVC), while ventricles expanded on average by 12.6+/-13.7% (as measured by PVVC).

There was a significant increase of total lesion volume (Table 2), 69.3% of which was due to expansion of chronic lesions and 30.7%-to new (confluent and free-standing) lesional activity.

**Table 2.** Longitudinal disability and lesion volume data.

|  | baseline | Follow-up | P value |
| --- | --- | --- | --- |
| EDSS Median values (range) | 1 (0-5) | 1 (0-6.5) |  |
| Total lesion volume | 5641+/-5033 | 6385+/-5424 | <0.001 |
| Chronic lesion volume | 5641+/-5033 | 6148+/-5383 | <0.001 |
| New lesional activity (confluent and free-standing lesions) |  | 237+/-235 |  |

Ventricular enlargement significantly correlated with EDSS at baseline (r^2^ =0.14, p=0.008) and EDSS change during study duration (r^2^ =0.16, p=0.006). Baseline EDSS (but not EDSS change) also correlated with change of brain volume (r^2^ =0.11, p=0.02)

The average increase in MD was significantly larger in new free-standing lesions than in the expanding component of chronic lesions (0.23+/-0.13 vs 0.10+/-0.04 respectively, p<0.001, t-test), suggesting that acute inflammation produces more severe lesional damage compared to slow-burning inflammation at the rim of chronic lesions.

There was high degree of correlation between the volume of combined lesional activity (i.e. the sum of the volume of all three compartments) and the volume of ventricular enlargement (r^2^=0.67, p<0.001 or r^2^=0.68, p<0.001 for partial correlation adjusted for age, gender, disease duration and baseline lesion volume) (Fig. 1a). This association became considerably stronger when lesion volume was replaced with PVSI (i.e. lesion volume adjusted by the degree of tissue damage severity in each of the three lesional compartments), reaching r^2^=0.81, p<0.001 (Fig. 1b) (unchanged for partial correlation, r^2^=0.81, p<0.001 for partial correlation adjusted for age, gender, disease duration and baseline lesion volume).

**Fig 1.**
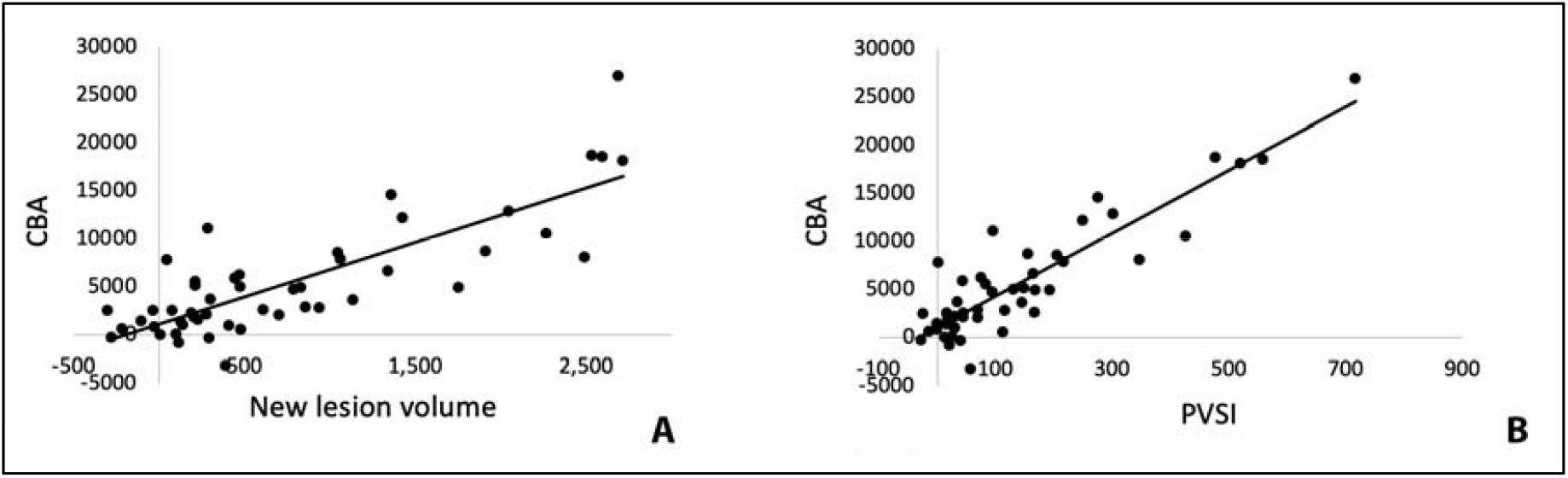
Correlation of CBA with new lesion volume (a) and PVSI (b).

The combined lesion activity volume also significantly (albeit to a much lesser degree) correlated with volume of whole-brain atrophy (r^2^=0.16, p=0.005 or r^2^=0.14, p=0.012 for partial correlation adjusted for age, gender, disease duration and baseline lesion volume) (Fig.2a), which improved after adjustment for the severity of tissue damage (r^2^=0.23, p<0.001 or r^2^=0.24, p=0.001 for partial correlation adjusted for age, gender, disease duration and baseline lesion volume) (Fig. 2b).

**Fig 2.**
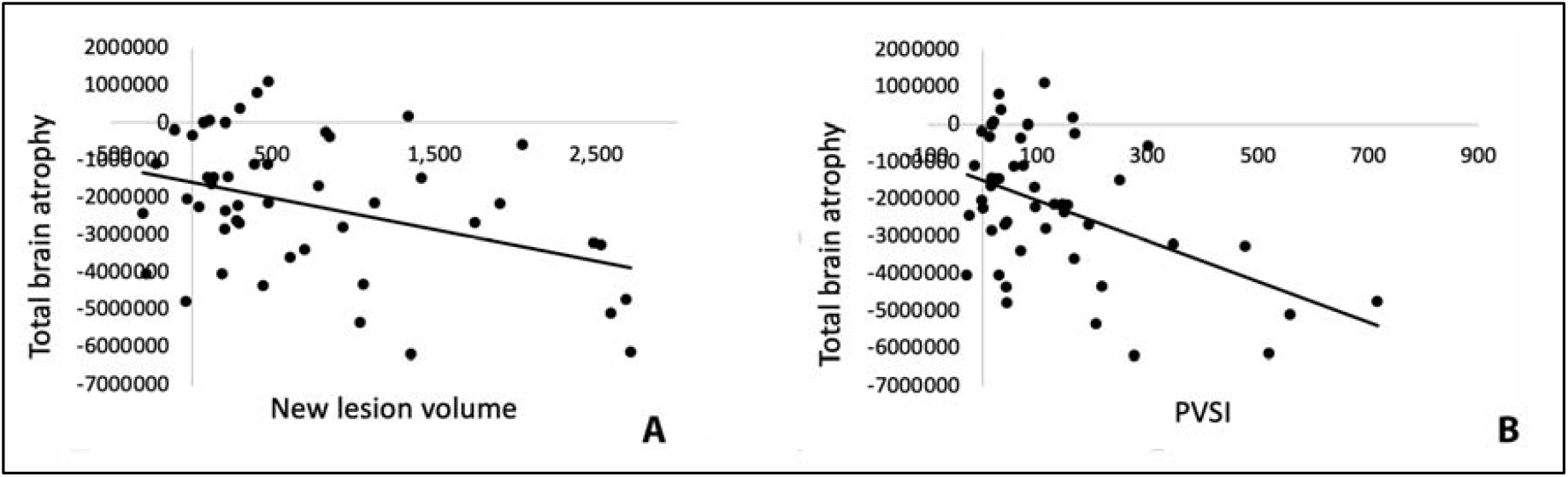
Correlation of volume of whole-brain tissue loss with new lesion volume (a) and PVSI (b).

Univariate linear regression analysis including all three compartments of lesion growth (i.e. expansion of chronic lesions, new confluent lesions and new free-standing lesions) adjusted for the severity of tissue damage in each compartment (model r^2^ =0.83) revealed that chronic lesion expansion was by far the largest contributor to ventricular enlargement (Standardized Coefficient Beta 0.68 (p<0.001) for expansion of chronic lesions compared to 0.29 (p=<0.001) for confluent lesions and 0.23 (p=0.001) for free-standing new lesions). When age, gender, disease duration, baseline lesion volume, baseline lesion MD, ventricular volume and treatment at the beginning of the study were added to the model, its power (r^2^) increased to 0.90 (Fig 3a).

**Fig. 3.**
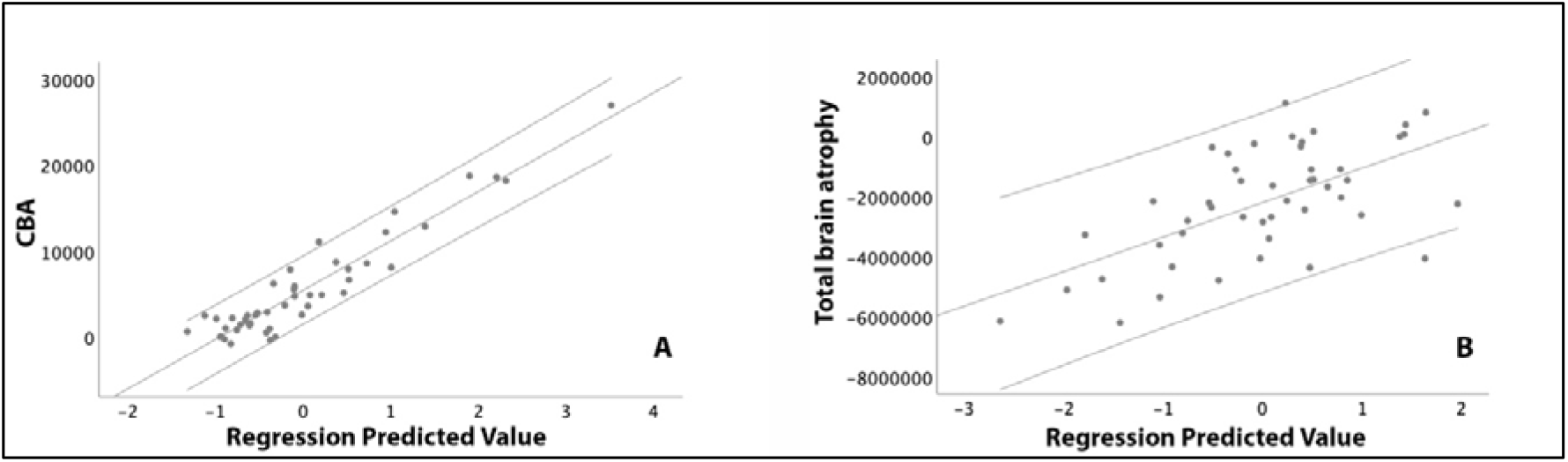
Linear regression model of CBA (a) and total brain atrophy (b).

PVSI in all three compartments remained highly significant contributors to the model (p<0.001 for all). Age and baseline ventricular volume also provided significant input to the model (p=0.045 and 0.015 respectively), while baseline lesion volume and MD in baseline lesions demonstrated borderline significance (p=0.06 for both).

The exclusion of 4 patients, who commenced fingolimod or natalizumab within 12 months of the final examination (to exclude the potential effect of pseudoatrophy) did not materially change the result (r^2^=0.86 or 0.28 for CBA and whole-brain atrophy using linear regression model).

Notably, volumetric parameters (i.e. lesion volume without adjustment for the severity of tissue damage) explained only 71% of CBA, measured by change in ventricular volume over the course of the study.

Similar relationships, but on a considerably lesser scale, were observed between lesional activity and whole-brain volume change. Linear regression analysis of the three components of lesion growth (model r^2^ =0.26) also showed that chronic lesion expansion was the largest contributor to whole-brain volume loss (Standardized Coefficient Beta 0.382 (p=0.035), 0.043 (p=0.8) and 0.284 (p=0.1) for expansion of chronic lesions, confluent and free-standing new lesions correspondingly). Adding age, gender, disease duration, baseline lesion volume, baseline lesion MD and treatment to the model increased its predictive power (r^2^=0.38) (Fig. 3b).

Again, using volumetric parameters only explained 31% of whole-brain tissue loss.

## Discussion

The measurement of brain atrophy is now widely used in neuroimaging and clinical studies of neurodegenerative diseases and has been employed as a biomarker of treatment efficacy in clinical trials, particularly in MS(Mak et al., 2017) (Zivadinov et al., 2016). Therefore, understanding mechanisms leading to brain atrophy are of a paramount significance. Ventricular enlargement has been suggested as potential surrogate marker of neurodegeneration(Kalkers et al., 2002) (Vrenken et al., 2014). It has significant advantage over conventional whole-brain measures of atrophy when personalized assessment, precision, computational speed, automated quantification and preferential central (periventricular lesion position are concerned (Zivadinov et al., 2016).

In the current study we examined the relationship of CBA, as measured by ventricular CSF enlargement, with progressive lesion-related tissue injury in patients with RRMS. We demonstrated that in RRMS patients combination of the new lesional activity, degree of tissue loss inside new lesional tissue and other demographic and MRI metrics explains 90% of variance in central brain atrophy. This association, driven primarily by chronic lesion expansion, was not affected by the potentially confounding effect of pseudoatrophy, reported in patients receiving particular DMTs (Vidal-Jordana et al., 2013) (Preziosa et al., 2020), since exclusion of patients who commenced fingolimod or natalizumab within 12 months of the final examination did not alter the result.

The remaining variability of CBA can, at least partially, be attributed to transients fluctuation of ventricular size (+/-6%), reported recently in MS patients(Millward et al., 2020).

### Severity of lesional tissue damage

To account for the severity of lesional tissue damage, we introduced the progressive volume/severity index (PVSI), which combines the volume of lesional tissue (new or chronic expanding) and an indicative measure of axonal loss (change in MD) within this region.

Considering potentially divergent pathomechanisms of acute and ‘slow burning’ inflammation in MS and a high likelihood that the severity of tissue damage in these zones differs, we calculated PVSI independently for each lesional compartment.

The addition of a measure of lesional tissue severity significantly improved interpretation of the nature of CBA, increasing the predictive value of lesional activity by >20%. Transected axons within lesions are largely replaced by gliosis(Andravizou et al., 2019), resulting in tissue rarefication and increased MD, but not lesion shrinking(S. A. Klistorner et al., 2020). The importance of axonal loss within new lesional tissue as a determinant of CBA is therefore likely to be related to Wallerian and retrograde degeneration of the extra-lesional part of the transected axons, which results in disintegration and elimination of axonal structure and associated myelin, rather than volumetric loss of lesional tissue *per se*. Given that the length of the extra-lesional part of transected axons often exceeds the intra-lesional component by several fold, often reaching a length of 10-15 centimetres(A. Klistorner et al., 2015), collapse of white matter caused by Wallerian and dying-back degeneration of those axons and, consequently, the volume of periventricular brain tissue loss, is likely to be highly proportional to the number of intra-lesional transected axons, as measured by an increase in MD.

Semi-quantitative gradation of the severity of tissue damage in MS lesions has previously been attempted using degree of T1 hypointensity, which was suggested to be associated with axonal density and widening of the extracellular space (van Waesberghe et al., 1999)(Barkhof & van Walderveen, 1999)(Vavasour et al., 2007). Incorporation of ranked T1 hypointensity did, in fact, demonstrate improved correlation of lesion burden with clinical and cognitive measures of disease progression in MS (Valizadeh et al., 2021)(Gautam Adusumilli et al., 2018) (Tam, Traboulsee, Riddehough, Sheikhzadeh, & Dkb Li, 2011) (Thaler et al., 2015) (Kocsis et al., 2021) (Poonawalla et al., 2010). The degree of T1 hypointensity was also used to estimate the level of progressive tissue damage inside chronic MS lesions in clinical trials (Elliott et al., 2019).

However, to our knowledge, detailed quantitative assessment of the severity of new lesional tissue damage by T1 hypointensity in combination with the volume of corresponding lesions has not been utilised in the assessment of brain atrophy.

### Expansion of chronic lesions is the major determinant of central brain atrophy

A leading contribution of new lesional activity to CBA is not surprising considering the largely periventricular positions of MS lesions and, as a result, preferential loss of periventricular white matter due to Wallerian and retrograde degeneration of transected axons in related fibre pathways. (Miller, Barkhof, Frank, Pakrker, & Thompson, 2002)(Kalkers et al., 2002).

Our study confirmed that the expansion of chronic lesions in patients with RRMS is a primary determinant of total increased T2 total lesion load (which aligns with earlier publications(S. A. Klistorner et al., 2020)(Elliott et al., 2019)). However, we also demonstrated that brain tissue loss associated with slow-burning inflammation at the edge of chronic lesions is a primary driver of CBA. Indeed, despite the fact that severity of tissue damage (as measured by MD change) was relatively low, linear regression modelling demonstrated that PVSI of the expanding edge of chronic lesions explains more central brain atrophy than the combination of new confluent and free-standing lesions.

The observation that tissue damage (as measured by MD change) within the expanding part of chronic lesions accumulates slowly in comparison to new free-standing lesions supports the notion that smouldering inflammation is considerably less destructive. However, the magnitude of the volume of chronic expanding lesion tissue (which constituted 69% of the total increase in lesion load) amplified its impact on brain atrophy. This finding is corroborates the earlier finding that, in MS patients receiving disease-modifying treatment, the expansion of chronic lesions is a better predictor of clinical progression and transition to secondary progressive disease than the formation of acute lesions(Elliott et al., 2019) (Absinta et al., 2019) (S. Klistorner et al., 2021)

Our observation that the severity of tissue damage in expanding and new lesions differs reinforces the previously suggestion that acute and smouldering inflammation have distinct pathomechanisms (Prineas et al., 2006)(Dal-Bianco et al., 2017) (Lassmann, 2007) (S. A. Klistorner et al., 2020). The former is driven by early damage to the blood–brain barrier (BBB) and recruitment of a substantial adaptive immune response that ultimately leads to inflammatory demyelination and axonal loss, while the latter has been attributed to compartmentalized slow-burning inflammation behind a relatively intact BBB that typically involves the lesion rim and is characterized histopathologically by the activation of CNS-resident cells (Geladaris, Häusler, & Weber, 2021) (Absinta, Sati, & Reich, 2016).

### Whole-brain atrophy

Whole-brain atrophy was notably less dependent on volume and severity of new lesional activity, which explained at most only about 1/3 of the change, suggesting that other factors, such as focal cortical lesions, grey matter atrophy due to retrograde degeneration and cortical neurone/synapse loss, oxidative injury of cortical neurons, meningeal inflammation or diffuse damage of the normal appearing white matter (NAWM) are also likely to contribute to whole-brain tissue loss (Haider et al., 2016) (Monaco, Nicholas, Reynolds, & Magliozzi, 2020) (Zivadinov et al., 2016) (Filippi et al., 2012) (Zhang et al., 2021). Furthermore, the smaller magnitude and higher variability of whole-brain atrophy are also likely to contribute to observed reduction in correlation.

### Limitations

This study has several limitations, including a relatively small sample size (50 subjects) with no direct control group and the fact that all but one patient was treated with DMTs, with potentially differential effects on lesion dynamics. Furthermore, 16 patients changed treatment category during the study period. However enrolling controls on no DMT is not feasible.

## Conclusion

In conclusion, our data suggest that CBA is almost entirely explained by the combination of the volume and severity of lesional tissue activity. Furthermore, the expansion of chronic lesions plays a central role in this process. The results of the study provide a better understanding of the mechanisms leading to loss of central brain tissue in patients with multiple sclerosis.

## Data Availability

All data produced in the present study are available upon reasonable request to the authors

